# A Systematic Review of the Protective Effect of Prior SARS-CoV-2 Infection on Repeat Infection

**DOI:** 10.1101/2021.08.27.21262741

**Authors:** N Kojima, NK Shrestha, JD Klausner

## Abstract

**Introduction:** We systematically reviewed studies to estimate the risk of SARS-CoV-2 reinfection among those previously infected with SARS-CoV-2.

**Methods:** For this systematic review, we searched scientific publications on PubMed and, the pre-print server, MedRxiv through August 18, 2021. Eligible studies were retrieved on August 18, 2021. We used the following search term on PubMed: (((“Cohort Studies”[Majr]) AND (“COVID-19”[Mesh] OR “SARS-CoV-2”[Mesh])) OR “Reinfection”[Majr]) OR “Reinfection”[Mesh]. We used the following search term on MedRxiv: “Cohort Studies” AND “COVID-19” OR “SARS-CoV-2” AND “Reinfection”. The search terms were broad to encompass all possibilities for applicable studies. There were no restrictions on the date of publication. Studies that did not describe cohorts with estimates of the risk of SARS-CoV-2 reinfection among those with previous infection were excluded. Studies that included vaccinated participants were either excluded or limited to sub-groups of non-vaccinated individuals. To identify relevant studies with appropriate control groups, we developed the following criteria for studies to be included in the systematic analysis: (1) baseline polymerase chain reaction (PCR) testing, (2) a negative comparison group, (3) longitudinal follow-up, (4) a cohort of human participants, i.e., not a case report or case series, and (5) outcome determined by PCR. The review was conducted following PRISMA guidelines. We assessed for selection, information, and analysis bias, per PRISMA guidelines.

**Results:** We identified 1,392 reports. Of those, 10 studies were eligible for our systematic review. The weighted average risk reduction against reinfection was 90.4% with a standard deviation of 7.7%. Protection against SARS-CoV-2 reinfection was observed for up to 10 months. Studies had potential information, selection, and analysis biases.

**Conclusions:** The protective effect of prior SARS-CoV-2 infection on re-infection is high and similar to the protective effect of vaccination. More research is needed to characterize the duration of protection and the impact of different SARS-CoV-2 variants.

## Introduction

Severe acute respiratory syndrome coronavirus 2 (SARS-CoV-2), the causative agent of coronavirus disease 2019 (COVID-19), is highly infectious and continues to cause substantial morbidity and mortality (Dong et al., 2020; Jin et al., 2020). Prior to the development of highly safe and effective vaccines for SARS-CoV-2 infection, scientists reported that a history of COVID-19 was associated with reduced risk of SARS-CoV-2 reinfection (Addetia et al., 2020). Virus-induced immunity has been described in many infections and is responsible for the decline of epidemic spread associated with exhaustion of the susceptible population (Rouse & Sehrawat, 2010). However, the duration and degree of the protective effect of SARS-CoV-2s-induced immunity is poorly studied.

Prior epidemiologic studies have found that individuals who are SARS-CoV-2 antibody-positive are protected against reinfection (Abu-Raddad et al., 2021; Harvey et al., 2021; Jeffery-Smith et al., 2021). Furthermore, investigators have reported that even those with prior SARS-CoV-2 infection *who lacked detectable antibodies* were at 80% lower risk of reinfection than people who were SARS-CoV-2 naïve (Breathnach et al., 2021). One retrospective study that analyzed test results among nearly 10,000 individuals with prior SARS-CoV-2 infection found that only 0.7% became reinfected with SARS-CoV-2 (Qureshi et al., 2021).

Other studies have also described reduced risk of infection, morbidity, and mortality among those with prior COVID-19. A study conducted in Austria found that the frequency of hospitalization and death due to a SARS-CoV-2 reinfection was 5 out of 14,840 (0.03%) and 1 out of 14,840 (0.01%), respectively (Pilz et al., 2021).

A history of COVID-19 may be as protective against reinfection as vaccination for SARS-CoV-2. A study investigating the frequency of repeat infection among laboratory personnel undergoing daily testing found no difference in SARS-CoV-2 infection rates between those with prior COVID-19 versus those vaccinated for SARS-CoV-2 infection (Kojima et al., 2021). Thompson et al. also reported that the decrease in risk of SARS-CoV-2 reinfection among those with prior infection was similar in magnitude to the relative risk reduction against SARS-CoV-2 infection among those who were vaccinated (Thompson et al., 2021). In a longitudinal study conducted among employees at the Cleveland Clinic, vaccination was not found to be associated with a lower risk of SARS-CoV-2 infection among people with prior COVID-19 (Shrestha et al., 2021).

Despite the availability of safe and effective vaccines, rates of SARS-CoV-2 infection are again increasing, especially among those without immunity (Christie et al., 2021). We aimed to determine the protective effect of previous infection among those who have not been also vaccinated against SARS-CoV-2. We systematically reviewed published longitudinal studies to estimate the risk of SARS-CoV-2 reinfection among those previously infected with SARS-CoV-2.

## Methods

For this systematic review, we searched scientific publications on PubMed and the pre-print server, MedRxiv. through August 18, 2021. Eligible studies were retrieved on August 18, 2021. We used the following search term on PubMed: (((“Cohort Studies”[Majr]) AND (“COVID-19”[Mesh] OR “SARS-CoV-2”[Mesh])) OR “Reinfection”[Majr]) OR “Reinfection”[Mesh]. We used the following search term on MedRxiv: “Cohort Studies” AND “COVID-19” OR “SARS-CoV-2” AND “Reinfection”. The search terms were broad to encompass all possibilities for applicable studies.

There were no restrictions on the date of publication. Studies that did not describe cohorts with estimates of the risk of SARS-CoV-2 reinfection among those with previous infection were excluded. Studies that included vaccinated participants were either excluded or limited to sub-groups of non-vaccinated individuals.

To identify relevant studies with appropriate control groups, we developed the following criteria for studies to be included in the systematic analysis: (1) baseline polymerase chain reaction (PCR) testing, (2) a negative comparison group, (3) longitudinal follow-up, (4) a cohort of human participants, i.e., not a case report or case series, and (5) outcome determined by PCR.

The review was conducted following PRISMA guidelines (Page et al., 2021). Two reviewers identified studies for the review. One reviewer collected data from each report and the other reviewer independently checked the work. Abstracts were reviewed and ineligible studies were not included. We reviewed selected reports to extract the following information: Author, year of publication, study cohort, reinfection risk, and follow-up time in person-years, when available. We assessed for selection, information, and analysis bias, per PRISMA guidelines. Due to the heterogeneity of the studies reviewed, sensitivity analyses and a meta-analysis was not attempted. Studies with outcomes were listed in a table.

There was no funding for this study.

## Results

We identified 1,392 reports (Figure). Of those reports, 10 studies met the above criteria from 6 different countries. The total population in the 10 studies included 9,930,470 individuals with a median observation period that ranged from one to 10.3 months.

**Figure.**
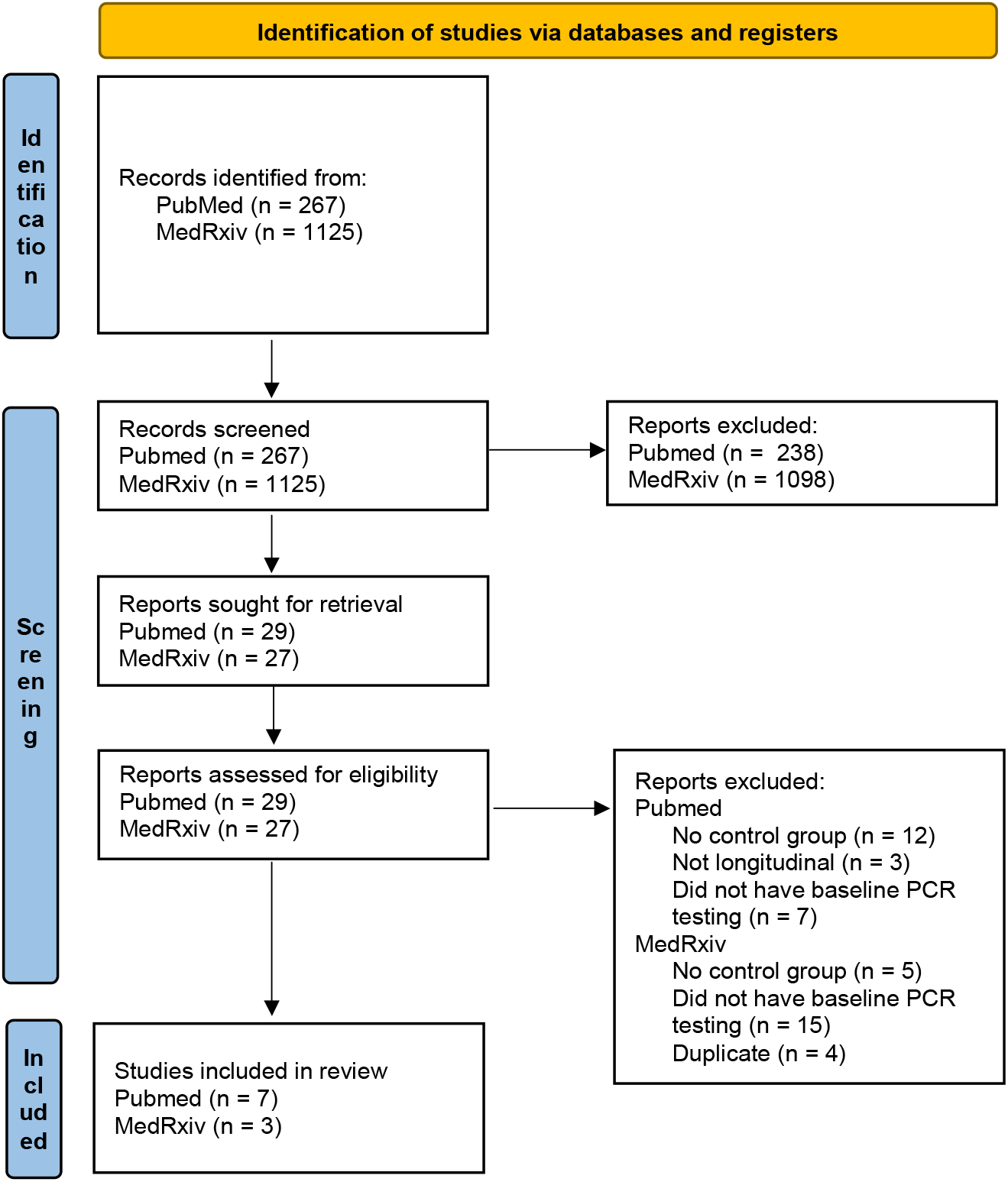
PRISMA flow diagram for systematic reviews

We found that the relative decreased risk of SARS-CoV-2 reinfection ranged between 80.5 to 100% compared to those without prior infection (Table). The weighted average risk reduction against reinfection was 90.4%, with a standard deviation of 7.7%.

**Table.**
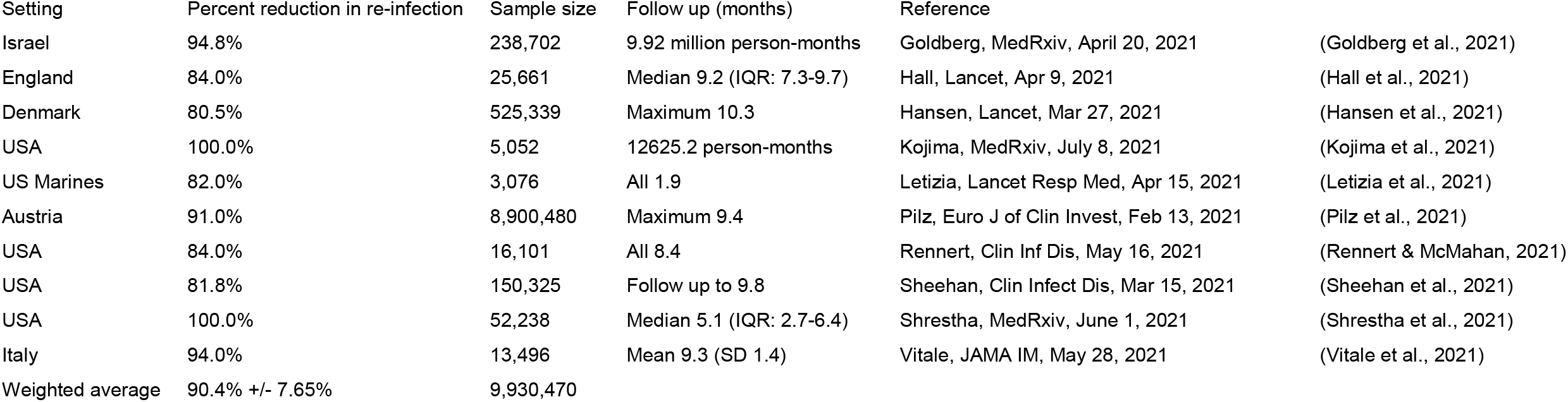
Studies through Aug 8, 2021 that show prior COVID-19 infection reduces risk for re-infection

The studies conducted by Goldberg et al., Hansen et al., Pilz et al., and Vitale et al., had cohorts compiled from national databases, which may have generated a selection bias towards people who had access to SARS-CoV-2 testing and were registered (Goldberg et al., 2021; Hansen et al., 2021; Pilz et al., 2021; Vitale et al., 2021). The study conducted by Hall et al. followed a cohort of 30,625 participants (Hall et al., 2021). In their study 51 participants withdrew and 4,913 participants were excluded because they did not have linked data for SARS-CoV-2 testing. That may have biased their study against people who did not have follow up testing, i.e., information and analysis bias. The study conducted by Letizia et al. studied young and healthy adults in the United States Marines who were undergoing basic training, and the housing conditions and interactions of individuals in that setting would not be readily extrapolated to the average population (Letizia et al., 2021). The study conducted by Rennert et al. studied university students that are younger and healthier than the average population (Rennert & McMahan, 2021). The study conducted by Shrestha et al. included younger and relatively healthier people, and since the study was done entirely after vaccines became available, there was also likely selection bias due to differential participation among those who decided not to get vaccinated (Shrestha et al., 2021).

The studies conducted by Hall et al., Rennert et al., and Sheehan et al., included follow-up that extended into the period when vaccines became available, and since vaccination was not controlled for, it is likely that vaccination among some subjects in the previously uninfected groups would have resulted in information bias resulting in an underestimation or overestimation of the effect size (Hall et al., 2021; Rennert & McMahan, 2021; Sheehan et al., 2021).

## Discussion

We systematically reviewed published longitudinal studies of SARS-CoV-2 reinfection with PCR confirmed initial and repeat infections. We found that the weighted average risk reduction against reinfection was 90.4%. Protection was observed up to 10 months. People with prior COVID-19 had a similar and durable level of protection when compared to those vaccinated against SARS-CoV-2 (Kojima et al., 2021; Stephens & McElrath, 2020).

In our systematic review, protection against SARS-CoV-2 reinfection was observed in up to 10 months following initial infection. It is not clear how long natural protection after infection will last. Biological studies have found persistent reservoirs of immunological active and antibody producing cells for up to 10 months or longer (Cohen et al., 2021).

The studies were conducted in 6 different countries. The studies ranged from participants that were younger than the national average (Letizia et al., 2021; Rennert & McMahan, 2021), as well as populations that were older than the national average (Vitale et al., 2021). Some studies followed participants at a national level (Goldberg et al., 2021; Hansen et al., 2021; Pilz et al., 2021), whereas other studies more closely followed cohorts (Letizia et al., 2021; Rennert & McMahan, 2021). While methodologies of studies differed, all reviewed studies consistently found decreased risk of reinfection among people with prior SARS-CoV-2 infection.

A recent United States Centers of Disease Control and Prevention (CDC) investigation conducted in Kentucky among persons with prior COVID-19 found that vaccination enhanced the protection of those with prior infection (Cavanaugh et al., 2021). In the CDC study, lack of vaccination after infection was associated with an increased odds of repeat SARS-CoV-2 infection, although the absolute increased risk of re-infection was very low. The study may have been biased due to different risk behaviors of the cases and controls. The study was not controlled for adherence with pandemic precautions (masking and social distancing), which would have been expected to have been different in the cases and controls.

Our study had several limitations. Our review was limited to studies with PCR confirmation of infection and re-infection. Multiple other studies, however, using SARS-CoV-2 antibody status as a measure of infection have similar results (Abu-Raddad et al., 2021; Harvey et al., 2021; A Leidi et al., 2021; A. Leidi et al., 2021). Our systematic review utilized some studies published on MedRxiv, a pre-print server. While MedRxiv had been helpful during the COVID-19 pandemic due to the rapid ability to disseminate information to colleagues, studies that were accessed on the site were not peer-reviewed. Furthermore, many of the studies cannot be replicated because they occurred in settings prior to the availability of vaccination against SARS-CoV-2 among people with history of infection.

Many of the studies including in our review followed people infected with SARS-CoV-2 earlier in the pandemic when infection was most likely with the original wildtype strain of SARS-CoV-2 before the development of variant strains. Therefore, our findings may differ in the current context of infections with exposure to variants that differ from the original infecting variant.

However a recent pre-print from a study conducted in the United Kingdom found among persons infected during a period of nearly exclusive Delta SARS-CoV-2 transmission, those fully vaccinated with BNT162b2 and ChAd0×1 (had similar levels of protection (82% and 67%, respectively) as those with previous infection (73%) (Pouwels; et al., 2021). Additionally, in a recent retrospective cohort study that was conducted in Israel which compared rates of SARS-CoV-2 infection or reinfection with the Delta variant among SARS-CoV-2-naïve individuals who received the BNT162b2 vaccine to people who had recovered from COVID-19, found that vaccinated, but SARS-CoV-2-naïve people, had an increased risk of infection with the Delta variant when compared to people who had recovered from COVID-19 (Gazit et al., 2021). This association was statistically significant in two models that either matched to the time of the first event (13.1-fold increased risk) or did not match to time of first event (6.0-fold increased risk).

## Implications

Our findings suggest that persons with prior COVID-19 are protected against SARS-CoV-2 reinfection. While protection has been observed in up to 10 months after initial infection, it is unclear how long the protection will last. Given the recency of new circulating variants like Delta, the protective effect of a previous infection with one variant and exposure to a different variant are uncertain. However, recent studies during the period of transmission of the Delta variant are promising.

## Conclusions

There is consistent epidemiologic evidence that prior SARS-CoV-2 infection provides substantial immunity to repeat SARS-CoV-2 infection. Prior SARS-CoV-2 infections provide similar protection when compared to vaccination for SARS-CoV-2. Longer follow-up studies are needed to determine how long protection lasts for natural immunity, especially among higher risk groups such as those with chronic medical conditions and those that are immunocompromised. More research is needed to investigate whether disease severity changes the risk of repeat infection. Finally, more research is needed to determine how much protection persists against emerging variants, like the Delta variant of SARS-CoV-2.

## Data Availability

This was conducted using publicly available data.

## Acknowledgements

None

## References

Abu-Raddad, L. J., Chemaitelly, H., Coyle, P., Malek, J. A., Ahmed, A. A., Mohamoud, Y. A., Younuskunju, S., Ayoub, H. H., Al Kanaani, Z., Al Kuwari, E., Butt, A. A., Jeremijenko, A., Kaleeckal, A. H., Latif, A. N., Shaik, R. M., Abdul Rahim, H. F., Nasrallah, G. K., Yassine, H. M., Al Kuwari, M. G., Al Romaihi, H. E., Al-Thani, M. H., Al Khal, A., & Bertollini, R. (2021). SARS-CoV-2 antibody-positivity protects against reinfection for at least seven months with 95% efficacy. EClinicalMedicine, 35, 100861. https://doi.org/10.1016/j.eclinm.2021.100861

Addetia, A., Crawford, K. H. D., Dingens, A., Zhu, H., Roychoudhury, P., Huang, M. L., Jerome, K. R., Bloom, J. D., & Greninger, A. L. (2020). Neutralizing Antibodies Correlate with Protection from SARS-CoV-2 in Humans during a Fishery Vessel Outbreak with a High Attack Rate. J Clin Microbiol, 58(11). https://doi.org/10.1128/JCM.02107-20

Breathnach, A. S., Duncan, C. J. A., Bouzidi, K. E., Hanrath, A. T., Payne, B. A. I., Randell, P. A., Habibi, M. S., Riley, P. A., Planche, T. D., Busby, J. S., Sudhanva, M., Pallett, S. J. C., & Kelleher, W. P. (2021). Prior COVID-19 protects against reinfection, even in the absence of detectable antibodies. J Infect, 83(2), 237–279. https://doi.org/10.1016/j.jinf.2021.05.024

Cavanaugh, A., Spicer, K., Thoroughman, D., Glick, C., & Winter, K. (2021). Reduced Risk of Reinfection with SARS-CoV-2 After COVID-19 Vaccination — Kentucky, May–June 2021. MMWR Morb Mortal Wkly Rep. https://doi.org/10.15585/mmwr.mm7032e1

Christie, A., Brooks, J. T., Hicks, L. A., Sauber-Schatz, E. K., Yoder, J. S., Honein, M. A., & Team, C. C.-R. (2021). Guidance for Implementing COVID-19 Prevention Strategies in the Context of Varying Community Transmission Levels and Vaccination Coverage. MMWR Morb Mortal Wkly Rep, 70(30), 1044–1047. https://doi.org/10.15585/mmwr.mm7030e2

Cohen, K. W., Linderman, S. L., Moodie, Z., Czartoski, J., Lai, L., Mantus, G., Norwood, C., Nyhoff, L. E., Edara, V. V., Floyd, K., De Rosa, S. C., Ahmed, H., Whaley, R., Patel, S. N., Prigmore, B., Lemos, M. P., Davis, C. W., Furth, S., O’Keefe, J. B., Gharpure, M. P., Gunisetty, S., Stephens, K., Antia, R., Zarnitsyna, V. I., Stephens, D. S., Edupuganti, S., Rouphael, N., Anderson, E. J., Mehta, A. K., Wrammert, J., Suthar, M. S., Ahmed, R., & McElrath, M. J. (2021). Longitudinal analysis shows durable and broad immune memory after SARS-CoV-2 infection with persisting antibody responses and memory B and T cells. Cell Rep Med, 2(7), 100354. https://doi.org/10.1016/j.xcrm.2021.100354

Dong, E., Du, H., & Gardner, L. (2020). An interactive web-based dashboard to track COVID-19 in real time. Lancet Infect Dis, 20(5), 533–534. https://doi.org/10.1016/S1473-3099(20)30120-1

Gazit, S., Shlezinger, R., Perez, G., Lotan, R., Peretz, A., Ben-Tov, A., Cohen, D., Muhsen, K., Chodick, G., & Patalon, T. (2021). Comparing SARS-CoV-2 natural immunity to vaccine-induced immunity: reinfections versus breakthrough infections. medRxiv. https://doi.org/10.1101/2021.08.24.21262415

Goldberg, Y., Mandel, M., Woodbridge, Y., Fluss, R., Novikov, I., Yaari, R., Ziv, A., Freedman, L., & Huppert, A. (2021). Protection of previous SARS-CoV-2 infection is similar to that of BNT162b2 vaccine protection: A three-month nationwide experience from Israel. medRxiv.

Hall, V. J., Foulkes, S., Charlett, A., Atti, A., Monk, E. J. M., Simmons, R., Wellington, E., Cole, M. J., Saei, A., Oguti, B., Munro, K., Wallace, S., Kirwan, P. D., Shrotri, M., Vusirikala, A., Rokadiya, S., Kall, M., Zambon, M., Ramsay, M., Brooks, T., Brown, C. S., Chand, M. A., Hopkins, S., & Group, S. S. (2021). SARS-CoV-2 infection rates of antibody-positive compared with antibody-negative health-care workers in England: a large, multicentre, prospective cohort study (SIREN). Lancet, 397(10283), 1459–1469. https://doi.org/10.1016/S0140-6736(21)00675-9

Hansen, C. H., Michlmayr, D., Gubbels, S. M., Molbak, K., & Ethelberg, S. (2021). Assessment of protection against reinfection with SARS-CoV-2 among 4 million PCR-tested individuals in Denmark in 2020: a population-level observational study. Lancet, 397(10280), 1204–1212. https://doi.org/10.1016/S0140-6736(21)00575-4

Harvey, R. A., Rassen, J. A., Kabelac, C. A., Turenne, W., Leonard, S., Klesh, R., Meyer, W. A., 3rd, Kaufman, H. W., Anderson, S., Cohen, O., Petkov, V. I., Cronin, K. A., Van Dyke, A. L., Lowy, D. R., Sharpless, N. E., & Penberthy, L. T. (2021). Association of SARS-CoV-2 Seropositive Antibody Test With Risk of Future Infection. JAMA Intern Med, 181(5), 672–679. https://doi.org/10.1001/jamainternmed.2021.0366

Jeffery-Smith, A., Iyanger, N., Williams, S. V., Chow, J. Y., Aiano, F., Hoschler, K., Lackenby, A., Ellis, J., Platt, S., Miah, S., Brown, K., Amirthalingam, G., Patel, M., Ramsay, M. E., Gopal, R., Charlett, A., Ladhani, S. N., & Zambon, M. (2021). Antibodies to SARS-CoV-2 protect against re-infection during outbreaks in care homes, September and October 2020. Euro Surveill, 26(5). https://doi.org/10.2807/1560-7917.ES.2021.26.5.2100092

Jin, H., Liu, J., Cui, M., & Lu, L. (2020). Novel coronavirus pneumonia emergency in Zhuhai: impact and challenges. J Hosp Infect, 104(4), 452–453. https://doi.org/10.1016/j.jhin.2020.02.005

Kojima, N., Roshani, A., Brobeck, M., Baca, A., & Klausner, J. D. (2021). Incidence of Severe Acute Respiratory Syndrome Coronavirus-2 infection among previously infected or vaccinated employees. medRxiv. https://doi.org/10.1101/2021.07.03.21259976

Leidi, A., Berner, A., Roxane, D., Dubos, R., Koegler, F., Piumatti, G., Vuilleumier, N., Kaiser, L., Balavoine, J. F., Trono, D., Pittet, D., Chappuis, F., Kherad, O., Courvoisier, D., Azman, A. S., Zaballa, M. E., Guessous, I., & Stringhini, S. (2021). Occupational risk of SARS-CoV-2 infection and reinfection during the second pandemic surge: a cohort study. medRxiv. https://doi.org/10.1101/2021.08.06.21261419

Leidi, A., Koegler, F., Dumont, R., Dubos, R., Zaballa, M. E., Piumatti, G., Coen, M., Berner, A., Darbellay Farhoumand, P., Vetter, P., Vuilleumier, N., Kaiser, L., Courvoisier, D., Azman, A. S., Guessous, I., Stringhini, S., & group, S. E.-P. s. (2021). Risk of reinfection after seroconversion to SARS-CoV-2: A population-based propensity-score matched cohort study. Clin Infect Dis. https://doi.org/10.1093/cid/ciab495

Letizia, A. G., Ge, Y., Vangeti, S., Goforth, C., Weir, D. L., Kuzmina, N. A., Balinsky, C. A., Chen, H. W., Ewing, D., Soares-Schanoski, A., George, M. C., Graham, W. D., Jones, F., Bharaj, P., Lizewski, R. A., Lizewski, S. E., Marayag, J., Marjanovic, N., Miller, C. M., Mofsowitz, S., Nair, V. D., Nunez, E., Parent, D. M., Porter, C. K., Santa Ana, E., Schilling, M., Stadlbauer, D., Sugiharto, V. A., Termini, M., Sun, P., Tracy, R. P., Krammer, F., Bukreyev, A., Ramos, I., & Sealfon, S. C. (2021). SARS-CoV-2 seropositivity and subsequent infection risk in healthy young adults: a prospective cohort study. Lancet Respir Med. https://doi.org/10.1016/S2213-2600(21)00158-2

Page, M. J., McKenzie, J. E., Bossuyt, P. M., Boutron, I., Hoffmann, T. C., Mulrow, C. D., Shamseer, L., Tetzlaff, J. M., Akl, E. A., Brennan, S. E., Chou, R., Glanville, J., Grimshaw, J. M., Hrobjartsson, A., Lalu, M. M., Li, T., Loder, E. W., Mayo-Wilson, E., McDonald, S., McGuinness, L. A., Stewart, L. A., Thomas, J., Tricco, A. C., Welch, V. A., Whiting, P., & Moher, D. (2021). The PRISMA 2020 statement: an updated guideline for reporting systematic reviews. BMJ, 372, n71. https://doi.org/10.1136/bmj.n71

Pilz, S., Chakeri, A., Ioannidis, J. P., Richter, L., Theiler-Schwetz, V., Trummer, C., Krause, R., & Allerberger, F. (2021). SARS-CoV-2 re-infection risk in Austria. Eur J Clin Invest, 51(4), e13520. https://doi.org/10.1111/eci.13520

Pouwels;, K. B., Pritchard;, E., Matthews;, P. C., Stoesser;, N., Eyre;, D. W., Vihta;, K.-D., House;, T., Hay;, J., Bell;, J. I., Newton;, J. N., Farrar;, J., Crook;, D., Cook;, D., Rourke;, E., Studley;, R., Peto;, T., Diamond;, I., Walker;, A. S., & Team, C.-I. S. (2021). Impact of Delta on viral burden and vaccine effectiveness against new SARS-CoV-2 infections in the UK Nuffield Department of Medicine.

Qureshi, A. I., Baskett, W. I., Huang, W., Lobanova, I., Naqvi, S. H., & Shyu, C. R. (2021). Re-infection with SARS-CoV-2 in Patients Undergoing Serial Laboratory Testing. Clin Infect Dis. https://doi.org/10.1093/cid/ciab345

Rennert, L., & McMahan, C. (2021). Risk of SARS-CoV-2 reinfection in a university student population. Clin Infect Dis. https://doi.org/10.1093/cid/ciab454

Rouse, B. T., & Sehrawat, S. (2010). Immunity and immunopathology to viruses: what decides the outcome? Nat Rev Immunol, 10(7), 514–526. https://doi.org/10.1038/nri2802

Sheehan, M. M., Reddy, A. J., & Rothberg, M. B. (2021). Reinfection Rates among Patients who Previously Tested Positive for COVID-19: a Retrospective Cohort Study. Clin Infect Dis. https://doi.org/10.1093/cid/ciab234

Shrestha, N. K., Burke, P. C., Nowacki, A. S., Terpeluk, P., & Gordon, S. M. (2021). Necessity of COVID-19 vaccination in previously infected individuals. medRxiv.

Stephens, D. S., & McElrath, M. J. (2020). COVID-19 and the Path to Immunity. JAMA, 324(13), 1279–1281. https://doi.org/10.1001/jama.2020.16656

Thompson, M. G., Burgess, J. L., Naleway, A. L., Tyner, H. L., Yoon, S. K., Meece, J., Olsho, L. E. W., Caban-Martinez, A. J., Fowlkes, A., Lutrick, K., Kuntz, J. L., Dunnigan, K., Odean, M. J., Hegmann, K. T., Stefanski, E., Edwards, L. J., Schaefer-Solle, N., Grant, L., Ellingson, K., Groom, H. C., Zunie, T., Thiese, M. S., Ivacic, L., Wesley, M. G., Lamberte, J. M., Sun, X., Smith, M. E., Phillips, A. L., Groover, K. D., Yoo, Y. M., Gerald, J., Brown, R. T., Herring, M. K., Joseph, G., Beitel, S., Morrill, T. C., Mak, J., Rivers, P., Harris, K. M., Hunt, D. R., Arvay, M. L., Kutty, P., Fry, A. M., & Gaglani, M. (2021). Interim Estimates of Vaccine Effectiveness of BNT162b2 and mRNA-1273 COVID-19 Vaccines in Preventing SARS-CoV-2 Infection Among Health Care Personnel, First Responders, and Other Essential and Frontline Workers - Eight U.S. Locations, December 2020-March 2021. MMWR Morb Mortal Wkly Rep, 70(13), 495–500. https://doi.org/10.15585/mmwr.mm7013e3

Vitale, J., Mumoli, N., Clerici, P., De Paschale, M., Evangelista, I., Cei, M., & Mazzone, A. (2021). Assessment of SARS-CoV-2 Reinfection 1 Year After Primary Infection in a Population in Lombardy, Italy. JAMA Intern Med. https://doi.org/10.1001/jamainternmed.2021.2959

